# Long-Term Persistence and Recovery of Short-Term Memory Loss in COVID-19 Survivors: A 3.5-Year Follow-Up Study

**DOI:** 10.1101/2025.03.10.25323656

**Authors:** Rakesh. K. Jha, Ravi K. Narayan, Sanjeev K. Paikra, Sanjib K. Ghosh, Ravi Kirti, Deependra. K. Rai, Ayan Banerjee, Asim Sarfaraz, Ashutosh Kumar

## Abstract

**Background:** Short-term memory loss is a common persistent symptom reported in recovered COVID-19 patients. This study evaluated its long-term persistence in a cohort of hospitalized COVID-19 patients following discharge.

**Materials and Methods:** A total of 207 hospitalized COVID-19 patients were followed up for three and a half years after discharge. They were interviewed via telephone and asked about symptoms indicative of short-term memory loss, such as increased forgetfulness or difficulty retaining recent information. A memory loss score was then created based on their responses. A time-based recovery trend was plotted, which served as the basis for modeling the further course of the illness.

Additionally, a retrospective analysis of hospital records, including clinical, laboratory, and treatment data for each patient, was conducted. The test data were statistically compared with an age, sex, and comorbidity-matched cohort that did not exhibit significant long COVID symptoms.

**Results:** Short-term memory loss was observed in approximately 11.5% (24/207) of patients. After three and a half years, 25% (6/24) had fully recovered (faster recovery group), 37.5% (9/24) showed improvement (gradual recovery group), and 29% (7/24) exhibited little to no progress (slow recovery group). Two patients with memory loss passed away during the study. Time trend analysis revealed significant symptom recovery over the study period (p < 0.001). Model predictions based on trend data estimate the maximum recovery period for the gradual recovery group to be 3.7 years. However, symptoms in the slow recovery group may persist for up to 14 years. Higher age and comorbidities were the strongest predictors of recovery speed. In contrast, patient sex, blood group, hospital stay duration, illness severity, and levels of inflammatory, metabolic, and thrombosis markers during hospitalization showed no significant impact (p > 0.05) compared to the COVID-19 survivors with no memory loss or other long COVID symptoms.

**Conclusion:** Significant recovery from memory loss can occur over time in COVID-19 survivors; however, symptoms persist in most patients even three and a half years after hospital discharge. A longer follow-up period may be necessary to assess the long-term trajectory of long COVID patients.

## Introduction

COVID-19 has significantly impacted the central nervous system ^1^. Approximately 20% of COVID-19 survivors experience long-term cognitive impairment ^2^, collectively termed brain fog ^3^. Short-term memory loss, a key feature of brain fog, has been reported in approximately 19% of recovered COVID-19 patients ^4^. However, long-term follow-up studies assessing the persistence of these symptoms remain scarce, and the underlying pathogenesis of memory loss in long COVID is not well understood.

The mechanisms behind cognitive symptoms in COVID-19 patients remain controversial. It is unclear whether these symptoms result from direct SARS-CoV-2 invasion of brain tissue or are a consequence of systemic inflammation. While cell-based and animal-model studies provide strong evidence of viral injury to brain cells ^5,6^, human studies have yet to confirm active viral replication within neurons. Instead, most studies detecting viral markers have identified them primarily in the vascular endothelium ^7^. In contrast, extensive inflammatory damage has been observed in both vascular as well as neural components of brain tissue ^8–10^.

One possible explanation for the rarity of direct viral injury is the low expression of the canonical SARS-CoV-2 entry receptor ACE2 in the human brain, except in the olfactory epithelium and neurovascular endothelium ^11^. Postmortem studies have demonstrated significant neurovascular endothelial damage, suggesting an indirect mechanism of injury ^8,9^. However, recent evidence indicates that SARS-CoV-2 may enter brain cells, including neurons, via neuropilin-1 (NRP1), a transmembrane glycoprotein with high expression in the human hippocampus—an area critical for memory function ^12,13^. This association suggests a possible link between NRP1-mediated viral entry and cognitive dysfunction in COVID-19 patients.

Additionally, recent studies have reported virus-induced endothelial barrier disruption of the brain’s microvasculature ^14,15^. However, while these findings contribute to our understanding of acute COVID-related brain injury, they do not fully explain the long-term persistence of cognitive dysfunction in some survivors. More recent studies suggest that blood-brain barrier (BBB) disruption in long COVID patients may result from sustained systemic inflammation, leading to localized neuroinflammation.

The inconsistency of evidence regarding these mechanisms has made it difficult to fully elucidate the pathogenesis of cognitive dysfunction in long COVID. Multiple mechanisms likely contribute to the persistence of these symptoms ^16^. Furthermore, animal model-based studies may not fully capture the complexity of long-term neuro-COVID.

Long-term follow-up studies are crucial for understanding the prolonged effects of COVID-19, as they provide real-world, case-by-case insights. However, studies specifically evaluating short-term memory loss in COVID-19 survivors over extended periods remain limited ^17^. Thus, it is not clear how long this symptom may persist and the chances of its resolution.

In this study, we followed a cohort of COVID-19 survivors for three and a half years after hospital discharge to assess the persistence of short-term memory loss. Additionally, we retrospectively analyzed clinical and laboratory data from their hospital stay to investigate potential contributing factors. Finally, we modeled time-based recovery trends to predict future recovery patterns.

## Materials and Methods

A total of 207 hospitalized patients of COVID-19 (Male: 137, Female: 70, Age range: 7-85 years) were followed up for three and half years post-discharge, between June 2021 and January 2025). These patients were diagnosed with COVID-19 based on the Real-Time PCR testing of the nasopharyngeal swabs and had the severity of illness decided as ‘moderate’ or ‘severe’ on clinical evaluation by a team of medical experts as per standard screening protocol necessitating their hospital admission (Table S1). A genomic sequence analysis of the randomly selected nasopharyngeal swabs indicated the dominance of the Alpha and later Delta strains of SARS-CoV-2 infection in the studied cohort of patients^18^. As the COVID-19 vaccines were yet to be introduced in the study region, the patients were considered to have no prior acquired immunity against the COVID-19 infection or severity of the symptoms.

Patients were verbally interviewed through a telephone call and asked about the symptoms confirming the loss of short-term memory, such as increased forgetfulness or problems with retaining a piece of recent information related to daily life activity or life events, and responses were recorded. A standard questionnaire was framed for recording the interview responses (Table S2). To facilitate an easy understanding of the queries and accurate responses, the patients were given brief verbal training over the phone, with examples of short-term memory loss (Table S3). A long-term COVID-19 illness was considered only if the symptom persisted for three months or longer post-hospital discharge as per World Health Organization (WHO) guidelines ^19^. The time of occurrence and period of persistence of memory loss and presence of any other neurological/psychiatric symptoms, including sleep loss or that related to other systems, were noted for each case.

At subsequent follow-ups, the patients were asked whether they had completely recovered, improved, remained persistent, or worsened their memory loss symptoms. A total of seven follow-ups were made at intervals of six months. At the first follow-up, the study participants were diagnosed with ‘short-term memory loss’ as a long COVID symptom based on the established criteria ^19^ ; annotated in the study as “Memory Loss (ML) patients”). The responses from the patients were uniformly graded based on the recovery status described by the patient, and a memory loss (ML) score was created in descending order from worst involvement to complete recovery from the symptoms (6=worst, 5-1= increasing levels of improvement, 0=complete recovery) (Table S4). A time-based recovery trend was plotted, which served as the basis for modeling the further course of the illness.

Additionally, a retrospective analysis of hospital records, including clinical, laboratory, and treatment data for each patient, was conducted. Universal standards were adopted for the measurement thresholds for reporting the laboratory parameters used in this study (Table S5). The test data were statistically compared with age, sex, and a comorbidity-matched cohort of COVID-19 survivors that did not exhibit long-term memory loss or any other long-COVID symptoms; annotated in the study as “Non-Memory Loss (NML) patients”.

### Statistical and data analysis

Descriptive statistics were calculated for the metric data. A one- or two-tailed t-test was conducted to compare the test and control groups, with significance set at p ≤ 0.05. To assess differences among test subgroups, ANOVA was performed, followed by Tukey’s Honestly Significant Difference (HSD) test for post hoc analysis. Pearson’s correlation and linear regression were used to examine the relationship between the ML score and laboratory parameters. Serial correlations were assessed using fixed effects models to evaluate changes in the ML score over the follow-up period. The Chi-square test was applied to analyze nominal data, with significance set at p ≤ 0.05. (If total events were less than 40 or zero events were reported in the contingency table, Yates correction and Fisher exact test, respectively, were used). All statistical analyses were conducted using R version 4.4.2 (Pile of Leaves) and the Online Social Science Statistics software (https://www.socscistatistics.com/).

### Ethics statement

An ethical clearance was received for this study from the Institute Ethics Committee of All India Institute of Medical Sciences, Patna, Bihar, India (Reference No.: AIIMS/PAT/IEC/2020/558).

## Results

Memory loss was observed in approximately 11.5% (24/207) of patients. Based on recovery patterns, the ML patients were categorized into three distinct recovery groups: Faster Recovery Group (FRG)—patients who achieved complete resolution of symptoms during the follow-up period; Gradual Recovery Group (GRG)—patients who exhibited significant symptom improvement over time; and Slow Recovery Group (SRG)—patients who showed minimal to no improvement.

At the end of the 3.5-year follow-up period, 25% (6/24) of patients fully recovered (FRG), 37.5% (9/24) demonstrated gradual improvement (GRG), and 29% (7/24) exhibited little to no progress (SRG). In the SRG group, two patients showed worsening memory loss between at 5^th^ and 6^th^ follow-ups, respectively, however, they improved on the subsequent follow-up. One of them had a recent diagnosis of hypertension and hyperlipidaemia, however, no specific reasons could be detected for the worsening of the symptoms in the other patient. Further, two patients with memory loss passed away due to terminal illness during the study period. Time trend analysis revealed a significant pattern of symptom recovery across the three groups (p < 0.001) (Fig. 1, Table 1). Statistical modeling predicted the estimated time for complete recovery as 2.7 years for FRG (y= -1.17143X + 3.17143, y-Memory Loss Score, x-Time period), 3.7 years for GRG (y = -1.77714X + 6.62, y-Memory Loss Score, x-Time period), while symptoms in the SRG group were projected to persist for up to 14 years (y = -0.44714X + 6.25714, y-Memory Loss Score, x-Time period) (Fig. 3).

**Table 1.** A study of serial correlation of fixed effects between Memory Loss (ML) Score and follow-up period.

| Fixed effects | Estimate | Std. Error | t value | P value |
| --- | --- | --- | --- | --- |
| (Intercept) | 5.38961 | 0.51549 | 10.46 | 5.99e-11 *** |
| Time | -0.50487 | 0.04378 | -11.53 | < 2e-16 *** |
| Correlation of Fixed Effects: (Intr) Time -0.340 |  |  |  |  |
| Significance codes: 0 '***' 0.001 '**' 0.01 '*' 0.05 |  |  |  |  |

**Figure 1.**
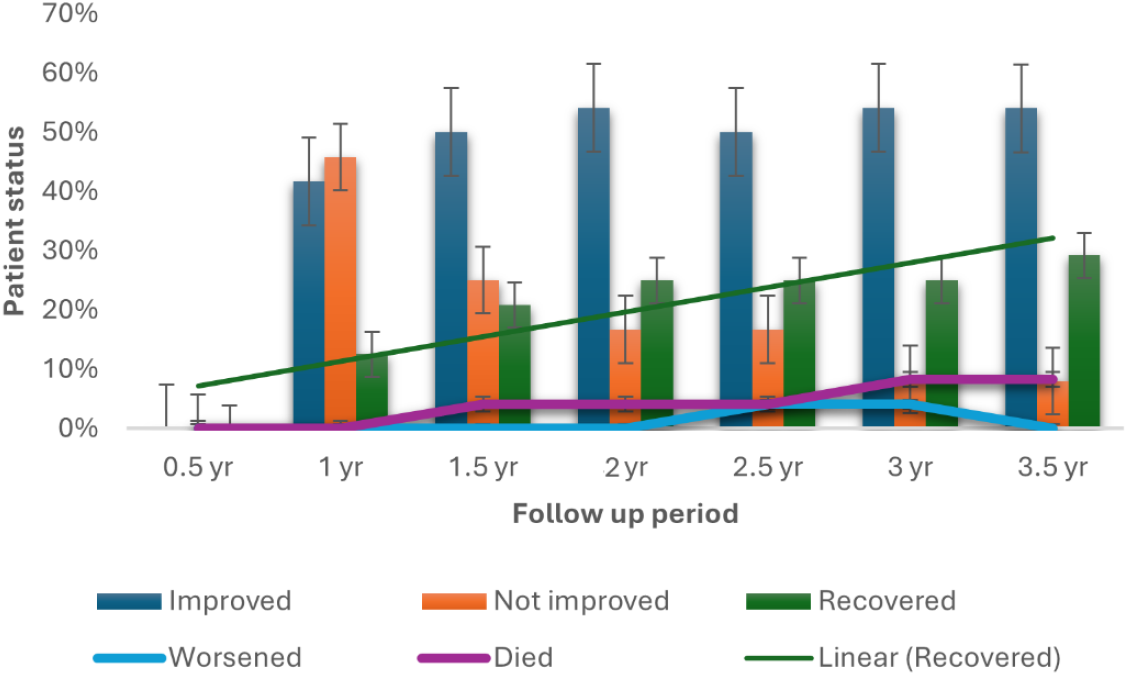
Time-based recovery trends of short-term memory loss in COVID-19 survivors over a 3.5-year follow-up period. The graph illustrates the proportion of patients in different recovery categories—Faster Recovery Group (FRG), Gradual Recovery Group (GRG), and Slow Recovery Group (SRG)—across multiple follow-up intervals. Statistical analysis revealed significant symptom resolution over time (p < 0.001).

**Figure 2.**
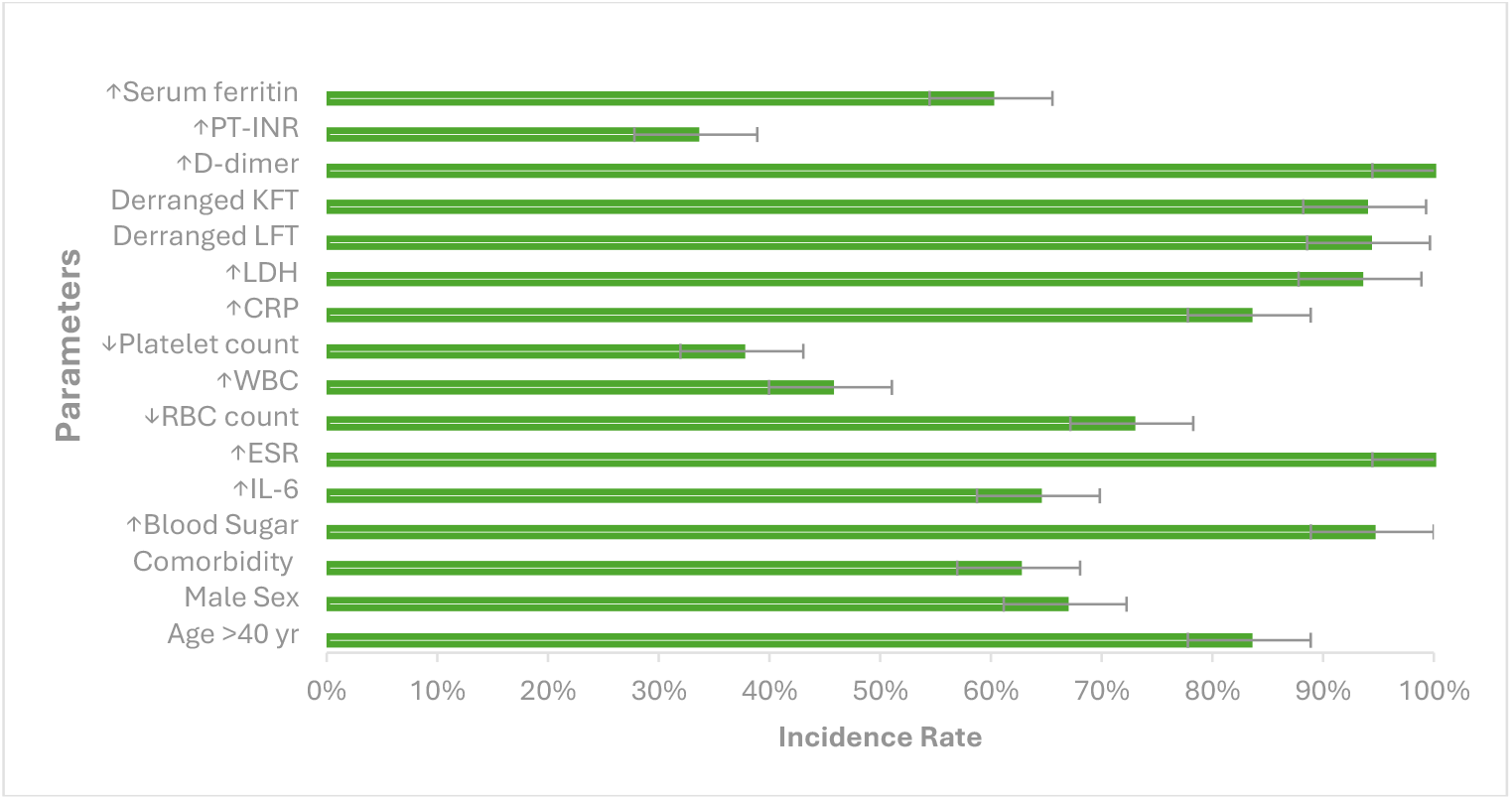
Incidence of key laboratory and clinical parameters in COVID-19 survivors with persistent short-term memory loss. Markers of systemic inflammation (IL-6, CRP, ESR, ferritin), metabolic dysregulation, and coagulation abnormalities were frequently observed but did not significantly differ from the COVID-19 survivors without short-term memory loss or other long COVID symptoms (p > 0.05).

**Figure 3.**
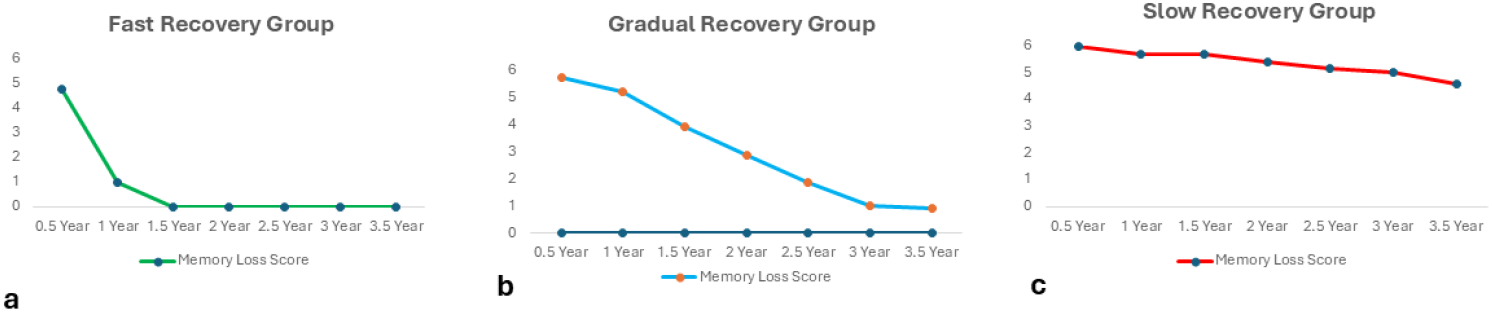
Projected recovery trajectories of short-term memory loss based on statistical modeling. Model equations indicate a progressive decline in memory loss scores over time (p < 0.001).

### Fixed Effects Model Analysis

The fixed effects model analysis of the Memory Loss (ML) scores (Table 1, Fig. 1) demonstrated a statistically significant association between time and symptom improvement. The intercept estimate (5.39, p < 0.001) (represents the baseline memory loss score), the time coefficient (-0.50, p < 0.001), and the negative correlation between time and intercept (- 0.340) further suggested that memory loss scores decreased progressively during the follow-up period.

### Predictors of Recovery

Higher age and comorbidities emerged as the most influential predictors of recovery speed. The ‘mean age’ of ML patients in the recovery groups were 48 years (range: 36-57 years) (FRG), 52 years (range: 28-72 years) (GRG), and 61 years (range: 46-68 years) (SRG), with a strong correlation between increased age and slower recovery ( r^2^ = 0.98) (Fig. 4).

**Figure 4.**
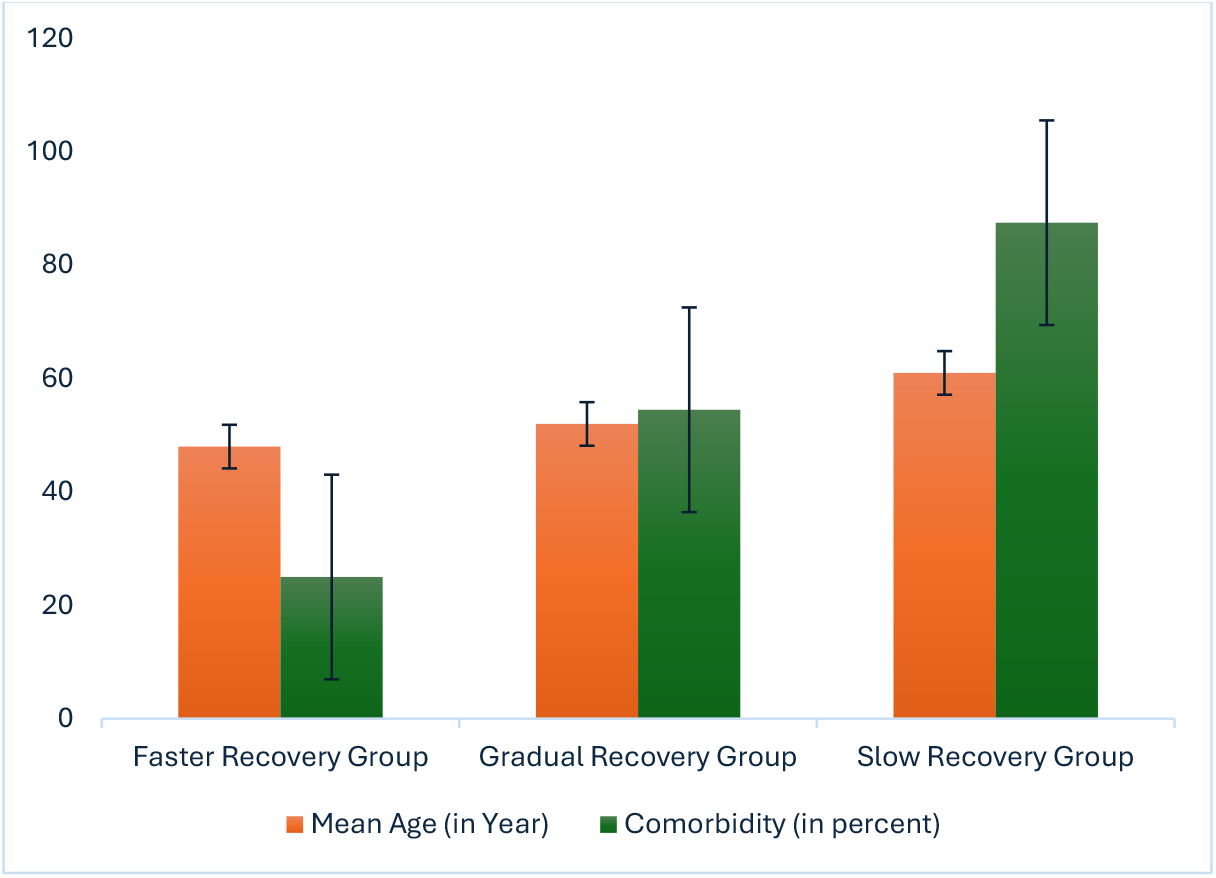
Mean age and comorbidity status of COVID-19 survivors with persistent short-term memory loss. A strong correlation was observed between increasing age and slower recovery (r^2^ = 0.98). Patients with comorbidities, such as diabetes and hypertension, demonstrated prolonged symptom persistence.

Additionally, the presence of comorbidities, such as diabetes, hypertension, or other systemic illnesses, was significantly associated with delayed recovery (FRG: 25%, GRG: 54.5%, SRG: 87.5%, r^2^ = 0.89).

### Associated Risk Factors

Patients in the test groups frequently exhibited several clinical and laboratory markers associated with memory loss (Figs. 2, 5). Higher age (>40 years), male sex, and the presence of comorbidities were commonly observed among affected individuals. Moreover, systemic inflammation markers—including elevated IL-6, ferritin, ESR, and CRP—were frequently detected. Coagulation abnormalities, such as increased PT/INR, elevated D-dimer levels, and reduced platelet count, were also prevalent. Additionally, metabolic disturbances (elevated blood sugar and LDH levels, decreased uric acid) and electrolyte imbalances (low sodium and calcium, high potassium) were noted in the ML patients. However, statistical comparisons with the NML patients did not yield significant differences (p > 0.05). No significant association with any blood group was found with the severity of the symptoms (p > 0.05). Associated neurological or psychiatric symptoms, such as loss of consciousness, anxiety, depression, or sleep loss, were very infrequent (< 1% of cases) and non-persistent in our studied cohort.

**Figure 5.**
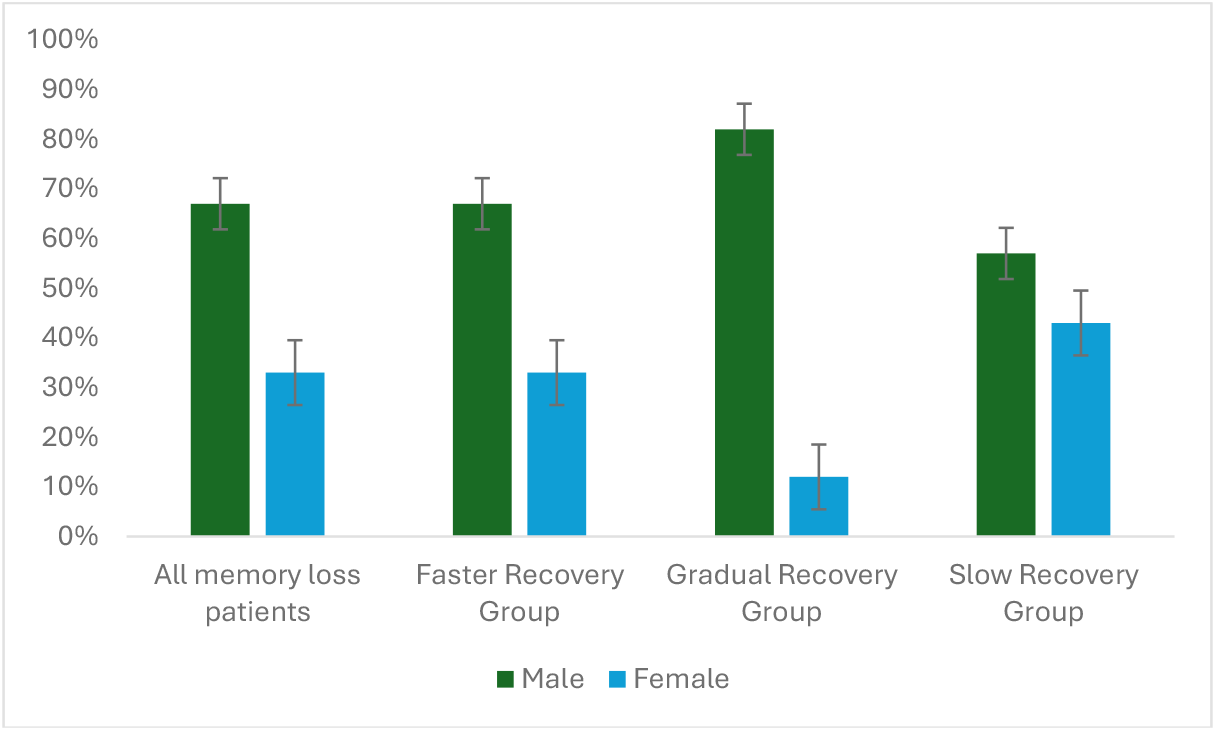
Sex distribution among COVID-19 survivors experiencing short-term memory loss. While both males and females were affected, a higher proportion of males exhibited memory impairment across all recovery categories. However, statistical analysis showed no significant sex-based differences in recovery rates (p > 0.05).

### Impact of Hospitalization and Illness Severity

The duration of hospital stay [ML patients: 11 (4-25) days; NML: 8 (2-18) days] and overall severity of illness [ML patients: Severe illness-1, Moderate illness-23, Mild illness: 0; NML patients: Severe illness-0, Moderate Illness-24, Mild Illness-0] did not significantly influence the recovery trajectory of memory loss when comparing the ML and NML cohorts (p > 0.05). Furthermore, no additional neurological or psychiatric symptoms were observed in patients diagnosed with memory loss throughout the study period.

## Discussion

Short-term memory loss has emerged as a persistent neurological concern among COVID-19 survivors. Our findings identified three distinct recovery trajectories, with 25% of patients achieving full recovery (FRG), 37.5% showing gradual improvement (GRG), and 29% experiencing persistent symptoms (SRG) over the 3.5-year follow-up period. Statistical modeling suggests that while FRG patients have recovered completely by 2.7 years (between the 5th and 6th follow-up), the GRG may recover fully in 3.7 years, which means they may be completely recovered by the next follow-up. However, the symptoms in SRG patients may persist for up to 14 years, indicating that complete recovery in this subset remains uncertain.

This study provides valuable insights into the long-term trajectory of post-COVID memory loss, offering optimism for many affected individuals. Given the limited long-term follow-up data available in the literature ^17^Our findings contribute to a growing body of evidence on the neurological consequences of COVID-19. Brief telephonic interviews using a structured questionnaire are a valuable and sensitive tool, yielding results comparable to in-person clinical examinations for assessing mild cognitive impairment^20^. They are particularly useful for evaluating patients in remote settings. Given the lack of standardized telephonic assessment tools for multi-stage evaluation of short-term memory loss, we developed a tool tailored to our study design and follow-up needs. The consistency of participant responses across seven follow-ups, as shown by the serial correlation analysis of fixed effects between memory loss scores and follow-up periods, highlights the tool’s reliability. While statistical modeling of memory loss persistence in gradual and slow recovery groups may not fully capture the actual recovery trajectory due to a limited sample size and intrinsic influencing factors, it reinforces the need for extended follow-up in long-COVID patients.

Age and comorbidities emerged as the most significant predictors of recovery speed, with older patients and those with underlying health conditions exhibiting slower recovery patterns. Although patients reported memory loss as a new onset symptom following COVID-19, an underlying influence of age-associated cognitive decline in our data, particularly in the higher age individuals, cannot be entirely ruled out. However, biological sex, illness severity, and hospital stay duration did not significantly impact recovery patterns. A retrospective analysis of laboratory data failed to identify a definitive systemic cause for the persistence or variability of memory loss, suggesting that other intrinsic patient-specific factors may play a role.

Existing literature proposes multiple mechanisms underlying cognitive impairment in COVID-19 survivors, including systemic inflammation, vascular thrombosis, metabolic imbalances, and direct viral invasion of brain cells ^16^. Short-term memory loss, primarily linked to hippocampal dysfunction, may result from impaired hippocampal neurogenesis ^21–23^ or disruption of the neurovascular endothelium, leading to blood-brain barrier (BBB) leakage ^24^. However, our retrospective analysis did not confirm systemic inflammation, vascular thrombosis, or metabolic disturbances as the primary drivers of memory loss, as these factors did not statistically differ from age- and comorbidity-matched NML patients. Nevertheless, their potential contribution to symptom development cannot be excluded.

The precise etiology of short-term memory loss in our patients remains uncertain. Given that cognitive dysfunction was not severely debilitating in most cases, direct neuronal injury seems unlikely as the primary mechanism. Instead, BBB disruption emerges as a more probable explanation, whether due to systemic factors or direct viral invasion ^25^. SARS-CoV-2’s affinity for ACE2-expressing vascular endothelial cells makes them highly susceptible to viral invasion ^11^. Greene et al. provided compelling evidence linking BBB impairment to long-COVID-associated cognitive dysfunction, showing that patients with brain fog exhibited BBB disruption on dynamic contrast-enhanced MRI and dysregulation of coagulation pathways in peripheral blood mononuclear cells. Increased endothelial adhesion of immune cells and inflammatory marker expression further support this hypothesis ^26^.

A compromised BBB may activate microglia and trigger neuroinflammatory cascades, leading to hippocampal dysfunction and persistent memory impairment ^27^. Another potential mechanism involves the direct invasion of hippocampal neurons by SARS-CoV-2, facilitated by alternative receptors such as neuropilin-1 (NRP1), which has been shown to mediate ACE2-dependent viral entry ^12,13^. Our recent research indicated high hippocampal expression of NRP1 ^13^ aligning with studies reporting persistent astrocytic and neuronal injury markers in the serum of long-COVID patients experiencing cognitive dysfunction ^12,28^.

Despite mounting evidence of BBB disruption and neuroinflammation in long COVID patients, definitive proof of direct SARS-CoV-2-induced neuronal injury remains lacking in the literature. It is possible that observed neuronal and astrocytic damage results from sustained systemic inflammation rather than direct viral assault ^26,29^. The ongoing debate between inflammatory-mediated damage and direct viral invasion warrants further investigation.

## Limitations

Our study provides essential long-term follow-up data on the trajectory of short-term memory loss indicative of cognitive impairment as a long-COVID symptom. However, several limitations must be considered. Reliance on self-reported telephone interviews may have introduced recall bias, and the lack of neuroimaging or biomarker analysis limits insights into hippocampal integrity and neuroinflammation. The potential effects of aging, psychological stress, and medications were not thoroughly evaluated. The study’s focus on hospitalized moderate-to-severe cases from a single institution restricts generalizability to milder cases. Moreover, we didn’t assess the impact of the specific strains of SARS-CoV-2 virus infection or role of intrinsic genetic factors in our studied cohort, which could have differentially affected the long COVID symptoms in the survivors ^30,31^. Additionally, as the cohort was unvaccinated at infection, the role of vaccination in cognitive recovery remains unclear, and reinfections were not systematically tracked. Fixed-interval follow-ups may have overlooked symptom fluctuations. Future research should include objective cognitive testing, neuroimaging, healthy controls, and broader samples to enhance validity and generalizability.

### Conclusion

Our study suggests that most long-COVID patients with short-term memory loss experience gradual improvement, with many achieving complete recovery over time. However, some patients exhibit persistent symptoms, necessitating longer follow-up studies to assess the full trajectory of cognitive recovery. We did not identify a direct systemic cause for memory loss in our cohort, instead proposing that a combination of systemic factors, intrinsic brain mechanisms, and localized hippocampal dysfunction may underlie these symptoms. Future studies utilizing advanced imaging and molecular analysis are required to elucidate the precise pathogenesis of cognitive impairment in long COVID.

## Data Availability

All data produced in the present study are available upon reasonable request to the authors.

## Acknowledgements

Funding for this study was provided by All India Institute of Medical Sciences-Patna.

## Author contributions

A.K., R.K.N., and R.K.J. conceptualized the study. R.K.J. conducted data collection. Data analysis was performed by R.K.J., A.K., and S.K.P., who also drafted the initial manuscript. S.K.G., R.K., D.K.R., A.B., and A.S. critically reviewed and finalized the manuscript. All authors approved the final version.

