## Supplementary Tables S1-5 for "Long-Term Persistence and Recovery of Short-Term Memory Loss in COVID-19 Survivors: A 3.5-Year Follow-Up Study"

**Supplementary Data:**

**Table S1. COVID-19 severity determination.***

| |
|---|
| **Mild**: upper respiratory tract symptoms and/or fever without shortness of breath or hypoxia) |
| **Moderate**: any of the following: 1. Respiratory rate > 24/min with breathlessness, 2. $SpO_2$ between 90% and <93% on room air) |
| **Severe:** any of the following: 1. Respiratory rate >30/min with breathlessness, 2. $SpO_2$ <90% on room air). |

*These classifications were based on the clinical criteria established by the Ministry of Health, Government of India (https://www.mohfw.gov.in/pdf/ClinicalManagementProtocolforCOVID19.pdf).

**Table S2. Questionnaire for post-survival assessment in discharged patients of COVID-19.**

**# At first Follow-up**

1. Have you experienced difficulties with memory or recall after being discharged?
   a. If yes, since when and for how many days
   b. Can you describe your symptoms
   c. Any other associated symptoms

2. Have you experienced a fever after being discharged?
   a. If yes, since when and for how many days
   b. Temperature observed – give measurements
   c. Any other associated symptoms
3. Have you experienced a cough after being discharged?
   a. If yes, since when and for how many days
   b. Any medication taken
   c. Any other associated symptoms
4. Have you observed any rash/discoloration of fingers/ toes after being discharged?
   a. If yes, since when and for how many days
   b. Any medication taken
   c. Any other associated symptoms
5. Have you had any difficulty or loss of sense of smell and taste after being discharged?
   a. If yes, since when and for how many days
   b. Any medication taken
   c. Any other associated symptoms
6. Have you had any difficulty in vision after being discharged?
   a. If yes, since when and for how many days
   b. Any medication taken

    c. Any other associated symptoms
7. Have you had any difficulty concentrating after being discharged?
    a. If yes, since when and for how many days
    b. Any medication taken
    c. Any other associated symptoms
8. Have you experienced any dizziness or loss of consciousness after being discharged?
    a. If yes, since when and for how many days
    b. Any medication taken
    c. Any other associated symptoms
9. Have you experienced an episode of seizure after being discharged?
    a. If yes, since when and for how many days
    b. Any medication taken
    c. Any other associated symptoms
10. Have you experienced any difficulty in speaking or walking after being discharged?
    a. If yes, since when and for how many days
    b. Any medication taken
    c. Any other associated symptoms
11. Have you experienced any difficulty in breathing or shortness of breath after being discharged?
    a. If yes, since when and for how many days
    b. Any medication taken
    c. Any other associated symptoms
12. Have you experienced any congestion in nose or runny nose after being discharged?
    a. If yes, since when and for how many days
    b. Any medication taken
    c. Any other associated symptoms
13. Have you experienced any chest pain/ compressions after being discharged?
    a. If yes, since when and for how many days
    b. Any medication taken
    c. Any other associated symptoms
14. Have you experienced nausea after being discharged?
    a. If yes, since when and for how many days
    b. Any medication taken
    c. Any other associated symptoms
15. Have you experienced any abdominal pain after being discharged?
    a. If yes, since when and for how many days
    b. Any medication taken
    c. Any other associated symptoms
16. Have you experienced an episode of diarrhoea after being discharged?
    a. If yes, since when and for how many days
    b. Any medication taken
    c. Any other associated symptoms
17. Have you experienced any urge to urinate after being discharged?
    a. If yes, since when and for how many days
    b. Any medication taken
    c. Any other associated symptoms
18. Have you experienced any body aches or pain after being discharged?
    a. If yes, since when and for how many days
    b. Any medication taken
    c. Any other associated symptoms

19. Have you experienced tiredness after being discharged?
    a. If yes, since when and for how many days
    b. Any medication taken
    c. Any other associated symptoms
20. Have you experienced loss of sleep after being discharged?
    a. If yes, since when and for how many days
    b. Any medication taken
    c. Any other associated symptoms

**# At Subsequent Follow-ups**

1. Did your memory loss symptoms
    a. Improved (better than before)
    b. Not improved
    c. Worsened (worse than before)
    d. Completely recovered
2. Did you develop any other neurological or psychiatric symptoms, or that related to any other physiological system since the last follow-up?
    a. If yes, since when and for how many days
    b. Any medication taken for this
    c. Any other associated symptoms
3. Were you diagnosed with Diabetes, Hypertension, or any other systemic illness since the last follow-up?
    a. If yes, since when and for how many days
    b. Who made the diagnosis
    c. Were any laboratory tests taken to confirm the diagnosis
    d. Any medications prescribed

**Table S3. Examples of short-term memory loss for brief training of the study participants.**

| Category | Example |
|---|---|
| **Daily Activities** | Forgetting why you walked into a room |
| **Conversations** | Asking the same question repeatedly |
| **Appointments** | Forgetting a meeting scheduled earlier in the day |
| **Objects** | Misplacing keys, glasses, or a phone and not recalling where you last had them |
| **Tasks** | Starting a chore but forgetting to finish it |
| **Names & Faces** | Meeting someone new and forgetting their name within minutes |
| **Directions** | Forgetting the route to a familiar place shortly after learning it |
| **Recent Events** | Forgetting what you ate for breakfast or a recent conversation |
| **Instructions** | Reading or hearing instructions but forgetting them almost immediately |
| **Phone Usage** | Unlocking the phone but forgetting why you picked it up |

**Table S4. Memory Loss (ML) Score based on the patient's recovery status.**

| ML Score | Recovery Status | Remarks |
|---|---|---|
| **6** | Worst involvement (severe memory loss, no improvement) | Completely impaired |
| **5** | Slight improvement, but still significant memory issues | Struggling heavily |
| **4** | Noticeable improvement, but frequent memory lapses | Forgetful often |
| **3** | Moderate improvement, occasional memory lapses | Somewhat better |
| **2** | Significant improvement, rare memory issues | Nearly recovered |
| **1** | Minimal symptoms, almost back to normal | Mild residual effects |
| **0** | Complete recovery, no memory issues | Fully recovered |

**Table S5. Laboratory parameters used in the study.**

| Laboratory Parameter | Threshold Value |
|---|---|
| **Metabolic Markers** | |
| Blood sugar (Fasting) | 75-110 mg/dl |
| Blood sugar (Post-prandial) | <140 mg/dl |
| Lactate dehydrogenase (LDH) | 230-460 U/L |
| Uric acid | 3.5-7.2 mg/dl |
| Sodium (Na) | 135-145 meq/L |
| Calcium (Ca) | 8.6-10 mg/dl |
| Potassium (K) | 3.5-5 meq/L |
| Chloride (Cl) | 98-107 meq/L |
| **Inflammatory Markers** | |
| IL-6 | <4.4 pg/ml |
| Ferritin | 10-291 ng/ml |
| Erythrocyte sedimentation rate (ESR) | 0-10 mm/hr |
| C-reactive protein (CRP) | 0-5 mg/L |
| **Thrombosis Markers** | |
| Prothrombin time (PT) | 11-16 s |
| International Normalized Ratio (INR) | 0.8-1 s |
| D-dimer | <0.2 µg/ml |
| Platelets | 150-450 thousand/µl |
